# A Protocol for Adapting a Mindfulness-Based Intervention with Contingency Management to Reduce Alcohol Misuse Among Latine Emerging Adults

**DOI:** 10.1101/2024.11.18.24315992

**Authors:** Michelle M Hospital, Sheila Alessi, Maria Eugenia Contreras Perez, Robbert Langwerden, Katherine R Perez, Jordan Quintana, Staci Leon Morris, Gus Castellanos, Sofia B Fernandez, Eric F Wagner

## Abstract

Emerging adulthood (ages 18-25 years old) is a critical period for the onset of problematic drinking, especially within underserved populations, such as Latine emerging adults. This protocol outlines the adaptation of a Mindfulness-Based Intervention (MBI) incorporating Contingency Management (CM) to address alcohol misuse in Latine young adults, a demographic with limited treatment access and increased vulnerability to substance use disorders. Grounded in Community-Engaged Research (CEnR) principles, this study employs a structured formative development approach to culturally and developmentally adapt a Mindfulness-Based Stress Reduction (MBSR) program. The intervention is designed to enhance emotional regulation and reduce alcohol misuse, supported by CM strategies to boost participant retention. This study’s objectives include assessing the feasibility, acceptability, and initial efficacy of the adapted MBI among non-treatment-seeking Latine emerging adults engaging in heavy episodic drinking. If successful, this protocol will provide a culturally resonant, non-traditional intervention model to mitigate alcohol misuse within this high-need population and inform future preventive efforts.

## Background

### Problematic drinking among Latine Emerging Adults

The transition from adolescence to adulthood (i.e., emerging adulthood, ages 18-25) is a critical period of role exploration, marked by the shift from dependence to independence, during which problematic alcohol and substance use peak. This period is characterized by increased experimentation with substances, influenced by the newfound autonomy and changing social environments of emerging adults (Schulenberg et al., 2004; Schulenberg & Maggs, 2015). Research indicates that college students, in particular, experience high levels of substance use as they navigate the challenges of this developmental stage, which includes managing academic pressures and social integration (Arnett, 2000; Gates et al., 2016).

Although Latine youth exhibit lower rates of heavy drinking compared to non-Latine Whites, they exhibit heightened risks of developing substance use disorders and negative outcomes (Alvarez et al., 2007; Cano et al., 2020; Mulia et al., 2009). Moreover, access to and utilization of treatment resources for alcohol misuse among Latine emerging adults lag significantly behind other racial/ethnic groups (Lui & Zamboanga, 2019; Vaeth et al., 2017), exacerbating existing health disparities. Barriers to seeking treatment, such as discrimination, cultural stigma, and misconceptions about treatment effectiveness, contribute to lower completion rates and further jeopardize health (NeMoyer et al., 2022). With the rapid growth of the Latino population in the United States (Funck & Lopez, 2022), addressing these barriers is essential to prevent widening health disparities (Ormiston et al., 2023). Developing interventions that specifically appeal to young Latine adults, who are less likely to seek and stay in traditional treatment, is crucial for improving health outcomes.

### Mindfulness-based interventions

Mindfulness-based interventions (MBIs) are a burgeoning nontraditional approach, to addressing health and mental health concerns, including alcohol misuse. If adapted to be culturally and developmentally sensitive, MBIs hold promise for reducing alcohol misuse among Latine emerging adults (Garland & Howard, 2018). Grounded in Buddhist philosophies and integrated into modern psychological practices, MBIs promote emotional self-regulation and present-moment awareness (Kabat-Zinn, 1982; Langer, 1989). Research indicates the potential of MBIs to interrupt processes associated with alcohol misuse, such as habituated cognitions and mindless behaviors (Bowen et al., 2014b; Ramadas et al., 2021). To our knowledge.

Research has shown that MBIs, such as mindfulness-based relapse prevention (MBRP), can significantly decrease the likelihood of relapse by helping individuals recovering from alcohol dependence manage cravings, reduce stress, and increase awareness of triggers (Bowen et al., 2014a, 2014b). However, there is a notable gap in research examining the application of MBIs for intervening in alcohol misuse, particularly heavy episodic drinking among young adults who have not yet escalated to alcohol dependence. This gap suggests a need for further exploration into how MBIs might be adapted to target this specific population before problematic drinking behaviors become more entrenched (Mermelstein & Garske, 2015). To our knowledge, this will be the first study to culturally and developmentally adapt an MBI to address problem drinking among non-treatment seeking Latine emerging adults.

#### Contingency Management to support MBI Retention

Despite the great promise of MBIs, retaining participants throughout the entirety of MBI treatment protocols remains quite difficult, particularly when working with individuals struggling with substance use, resulting in suboptimal outcomes (Nam & Toneatto, 2016). Adding a Contingency Management (CM) component to reinforce attendance at MBI sessions is an innovative approach that could support MBI retention. CM is a behavioral intervention that combines operant conditioning procedures and behavioral economic principles to motivate pro-health behaviors. It is highly effective at increasing abstinence (substance-free drug tests), attendance at treatment visits, and adherence with exercise and other health behaviors (Davis et al., 2016; DeFulio & Silverman, 2012; Lussier et al., 2006; Moore et al., 2015; Petry et al., 2015; Prendergast et al., 2006; Volpp, John, et al., 2008; Volpp, Loewenstein, et al., 2008; Weinstock et al., 2016)(Davis et al., 2016; DeFulio & Silverman, 2012; Lussier et al., 2006; Moore et al., 2015; Petry et al., 2015; Prendergast et al., 2006; Volpp, John, et al., 2008; Volpp, Loewenstein, et al., 2008; Weinstock et al., 2016).

Research on CM in the context of MBIs is emerging (Asfar et al., 2022; Carrico et al., 2018, 2019; Glasner et al., 2017; Minami et al., 2018, 2022; Peter et al., 2023). In a study among sexual minority men who used methamphetamine (N = 110; 42.7% Black/African American, 11.8% Latino), 12 weeks of CM for stimulant abstinence coupled with a positive affect and mindfulness intervention decreased self-reported stimulant use and craving (Carrico et al., 2018) through follow-up compared to CM only (Carrico et al., 2019). In a study among adults with stimulant use disorder (N = 63; 20.6% Hispanic, 44.4% African American, 71.4% male), among those with depressive disorder, 12 weeks of CM for stimulant abstinence plus mindfulness-based relapse prevention was associated with lower odds of stimulant use compared to CM with health education (Glasner et al., 2017). After demonstrating feasibility and acceptability, a randomized controlled trial (RCT) compared a smartphone-assisted mindfulness-based smoking cessation program plus CM for smoking abstinence to the enhanced standard smoking cessation condition (N = 49; 63.3% Latine, 30.6% Black, 75.5% female) (Minami et al., 2022) and found increased abstinence (biochemically determined) in the CM condition through the end-of-treatment. Thus, there is evidence that the addition of CM to a mindfulness program is feasible among racially and ethnically diverse samples. However, there remains a gap in our understanding of the benefit of pairing CM with an MBI focused on the reduction of alcohol misuse among Latine young adults who are not seeking treatment.

### Study Objectives

This study aims to introduce a culturally and developmentally adapted MBI, coupled with CM for promoting session attendance, to address alcohol misuse among non-treatment-seeking Latine emerging adults. By focusing on self-regulatory processes associated with alcohol misuse and leveraging the potential for culturally resonant aspects of mindfulness, this intervention seeks to offer a respectful, non-judgmental, and discrimination-free approach to alcohol misuse within this high-need population. The integration of CM aims to enhance participant engagement and retention, a critical factor given the historically high dropout rates in MBI trials targeting alcohol and substance use issues (Nam & Toneatto, 2016)

## Methods

### Research Design Overview

This study employs a structured formative development approach to create an adapted MBSR program tailored for Latine emerging adults (EAs) engaging in alcohol misuse. The integration of Community-Engaged Research (CeNR) principles aims to ensure developmental and cultural relevance (Task Force on the Principles of Community Engagement., 2011).

### Target Population and Eligibility

Participants will be Latine (self-identified) emerging adults (EAs), aged 18-25 years old, actively enrolled in a 4-year college program and engaging in heavy episodic drinking (HED) (defined as consuming 4/5 drinks for females/males within a single occasion at least twice in the preceding 30 days)(*Drinking Levels and Patterns Defined* | *National Institute on Alcohol Abuse and Alcoholism (NIAAA)*, n.d.). Exclusion criteria are five or more binge drinking episodes in the past 30 days, which averages more than once per week and weekly or more frequent use of illicit substances.

### Study Site

The study site is a prominent public university in South Florida, known for its substantial Latine student population and federal designation as a Hispanic Serving Institution (HSI). This setting provides an ideal environment for engaging with the target population to develop and implement the culturally adapted MBSR program.

### Stages of Adaptation

For this formative development of a developmentally and culturally acceptable adaptation of the MBSR intervention to reduce alcohol misuse among Latine EA, our primary outcomes are the acceptability and feasibility of the proposed program. Our design will involve an adaptation process consistent with CeNR principles. The study design draws from the National Center for Complementary and Integrative Health (NCCIH) Framework for Developing and Testing Mind and Body Interventions (*Research Framework* | *NCCIH*, n.d.) using an iterative method for Intervention Refinement and Optimization. This adaptation and refinement process will be guided by the five stages of cultural adaptations of behavioral health interventions (Barrera et al., 2013) (See Figure 1). Feedback will inform successive refinements of the MBSR curriculum, with pilot testing and subsequent revisions focused on maximizing cultural relevance and ecological validity.

**Figure 1.**
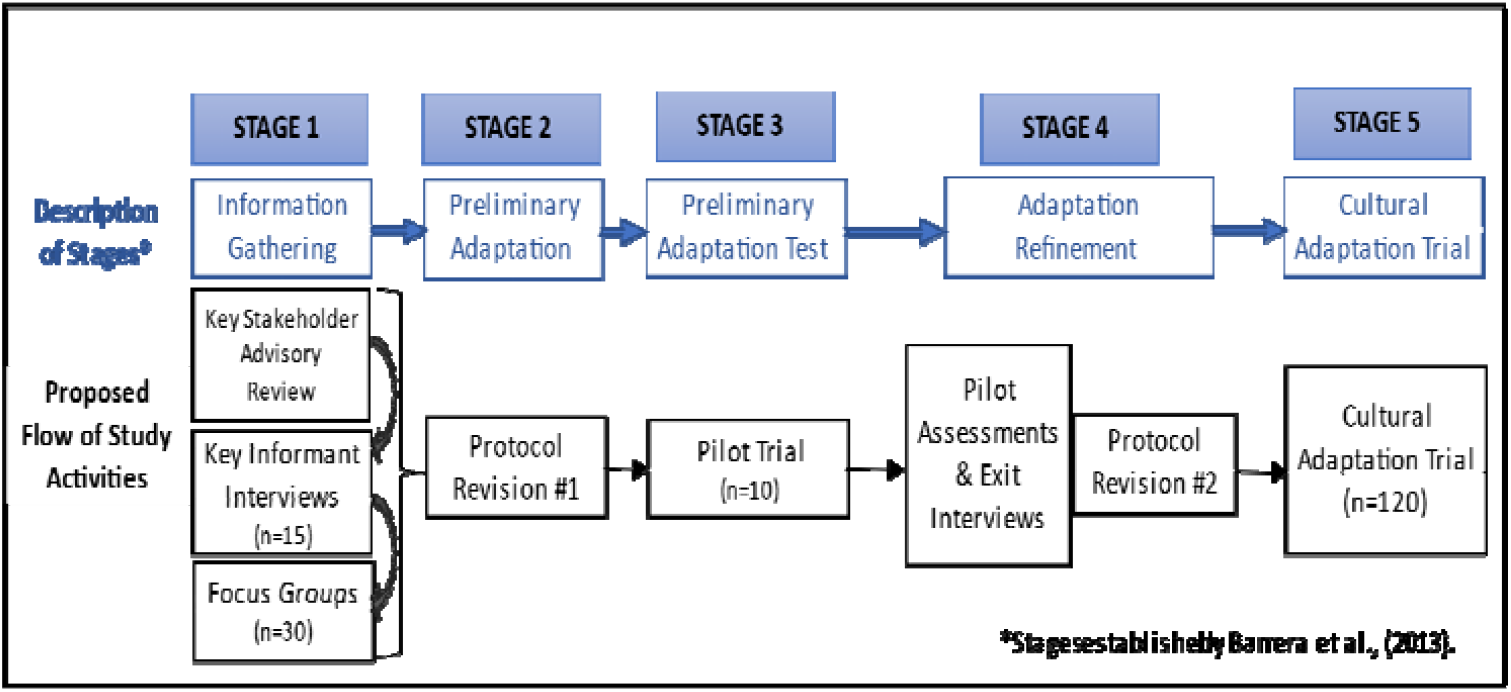
proposed stages of cultural adaptation.

### Stage 1: Information Gathering and Stakeholder Engagement

#### Key Stakeholder Advisory Review

Feedback will be initially sought from key stakeholders within the university community, including the Drug-Free Campus/Workplace Drug and Alcohol Abuse Taskforce (AODTF), comprised of faculty, students, staff, and administrators from various departments. The goal is to gather insights and assess face validity on the MBSR core elements and explore potential adaptations to enhance the program’s success in engaging the target population.

#### Key Informant Interviews

Latine EAs (n=15) engaging in alcohol misuse will be interviewed to understand their perspectives on mindfulness, potential benefits, barriers to participation, and how an MBSR intervention might address their stress, discrimination experiences, and motivations for drinking. These semi-structured interviews aim to inform adaptations to the recruitment, data collection, and intervention protocols.

#### Focus Groups

Following key informant interviews, focus groups with Latine EAs (n=30) will review the proposed adapted recruitment and data collection protocols as well as the MBSR curriculum to provide feedback on its relevance, utility, and acceptability. These discussions aim to refine the intervention, ensuring it resonates with the cultural, generational, and developmental contexts of the target population.

### Stage 2: Preliminary Adaptation

Data obtained from Stage 1 activities will form the foundation for operationalizing the adapted MBSR intervention manual and data collection protocols. Activities include refining assessment questions, developing recruitment materials and procedures, adapting MBSR session duration and activity sequence to enhance feasibility and relevance, creating intervention facilitator scripts, establishing CM procedures for sessions, and preparing training manuals for intervention and data collection staff.

### Stage 3: Pilot Trial

A pilot trial of the adapted 8-week (1 hour/week) MBSR program will be conducted with Latine EAs (n=10) engaging in alcohol misuse. Assessments will occur at two timepoints (baseline and post-intervention, see Figure 2). Additionally, post-intervention qualitative exit interviews will cover topics such as program structure, session content relevance and appeal, acceptability of CM protocols, data collection procedures, facilitator characteristics, barriers to participation, appropriateness of assessments, and overall impressions.

**Figure 2.**
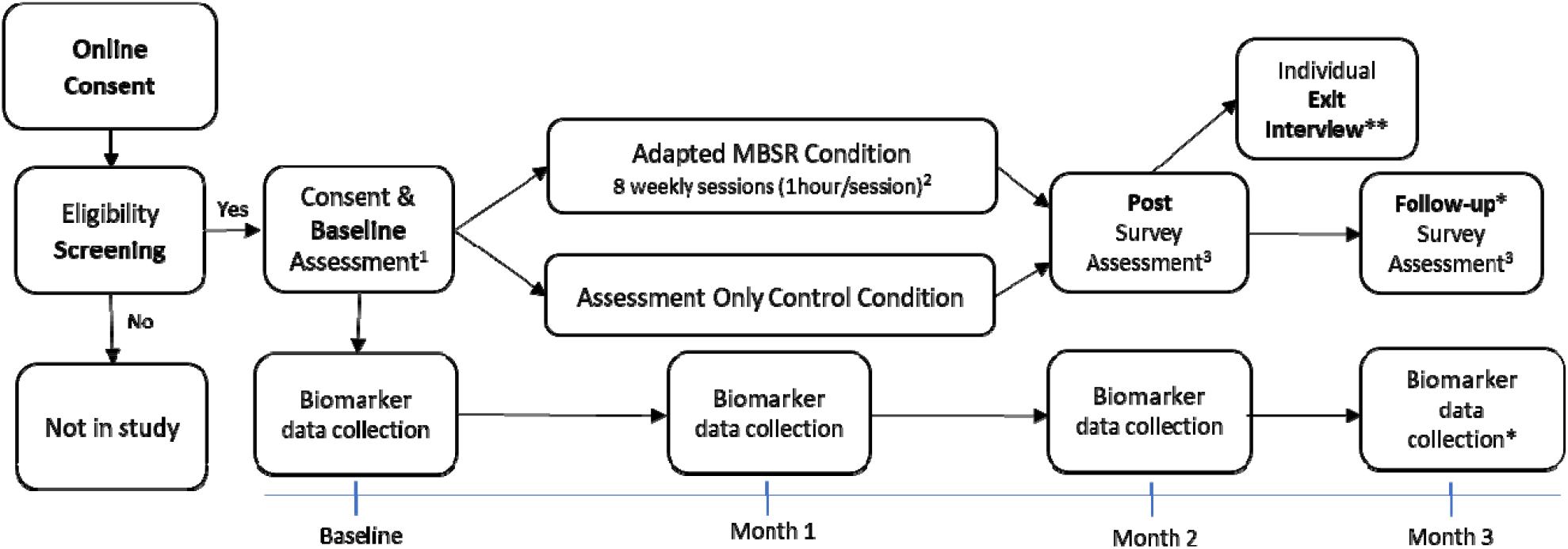
Participant Flow Chart Pilot trial (n=10) and Cultural Adaptation Trial (n=120) Notes:^1^ Participants will receive $50 for completion of Baseline survey assessment, ^2^ Participants in MBSR condition will earn draws and chances for prizes for session attendance. ^3^Participants will receive $25 each for completion of post and follow-up assessments. Participants will be paid $10 for each biomarker data collection visit. *Only applicable for Cultural Adaptation Trial participants

### Stage 4: Adaptation and Refinement

The project team, including an MBSR expert consultant, will engage in weekly meetings to use findings from the pilot trial to refine all protocols and materials. While essential components of the intervention (e.g., present moment awareness, decentering, and non-judgment) and the main topic areas will be maintained, revisions to the thematic content and sequence of activities/sessions will be considered based on the trial outcomes. The research team will conduct a thorough review of the pilot trial findings to finalize the MBSR intervention.

### Stage 5: Cultural Adaptation Trial

This study culminates in a Cultural Adaptation Trial to evaluate the efficacy and cultural appropriateness of the new adapted Mindfulness-Based Stress Reduction program for Latine emerging adults. Participants in the Cultural Adaptation Trial will be Latine emerging adults (N=120), meeting the same eligibility criteria as in the pilot trial.

### Recruitment and Randomization

For recruitment, we will leverage our prior Substance Abuse and Mental Health Services Administration (SAMHSA) funded-work in which we developed detailed protocols for on-campus, in-person student outreach and social media-based (e.g., Instagram) efforts in collaboration with the university’s Healthy Living Program, Counseling and Psychological Services and College of Public Health (Fernandez et al., 2019). To promote the study, we will leverage our experience conducting in-person outreach via information booths in highly visible and popular locations on campus as well as posting printed and digital flyers around campus. During the formative research, we will solicit information about potentially new social media platforms and explore other recruitment strategies.

### Assessment Schedule and Measures

The participant flow chart and assessment schedule are shown in Figure 2. A summary of the constructs and corresponding measures we plan to collect is in Table 1. Demographic data will include age, gender identity, sex at birth, race, nativity/generation status, years in the U.S., student status, household income, employment status, ethnicity and Latine heritage, partner status, sexual orientation, religious orientation, health insurance, as well as process measures of intervention satisfaction. Acceptability data will be collected for the Pilot and Cultural Adaptation Trials. Screening, baseline, and follow-up questionnaires will be completed online using a secure online data collection platform (e.g., REDCap) and are expected to require approximately 50-60 minutes for baseline and 30-40 minutes each for post and follow-up.

**Table 1.** Proposed Assessment Schedule (Pilot Trial and Cultural Adaptation Trial)

| **Construct** | **Baseline** | **Post** | **Follow-up\*** |
| --- | --- | --- | --- |
| Heavy Episodic Drinking (HED) | x | x | x |
| Alcohol consumption | x | x | x |
| Drinking Consequences | x | x | x |
| Drinking motives | x | x |  |
| Age of drinking onset | x |  |  |
| Heavy Episodic Drinking - alcohol biomarker phosphatidylethanol (PEth) | x | x | x |
| Other Substance use | x | x | x |
| Psychosocial stress | x | x | x |
| Stress (Heart Rate Variability) | x | x | x |
| Emotion Regulation | x | x | x |
| Impulsivity | x | x | x |
| Discrimination | x | x |  |
| Coping Strategies | x | x | x |
| Well-Being | x | x | x |
| Mindfulness | x | x | x |
| Depression symptoms | x | x | x |
| Anxiety symptoms | x | x | x |
| Suicidal Ideation | x | x | x |

\* Cultural Adaptation Trial participants will complete a Follow-up (3 months after baseline)

#### Biomarker data collection

The direct alcohol biomarker phosphatidylethanol (PEth) will be measured using dried blood spots (DBS) to assess for the presence/absence of episodes of Heavy Episodic Drinking during the prior 3-4 weeks (Piano et al., 2015). After a finger-prick, the coded DBS will be sent to US Drug Testing Laboratories (USDTL, Des Plaines, IL) for quantitative PEth analyses by liquid chromatography with tandem mass spectrometry. Non-medical personnel can readily be trained to collect and send specimens. Heart rate variability (HRV) is a physiological measure of stress and will be measured (Kim et al., 2018) using procedures similar to Chu and colleagues (Chu et al., 2017). Briefly, participants will be asked to sit and relax with no other tasks or stimulation for 5-10□minutes. During the last 5□minutes, heart rate data will be collected. Specific indices of HRV (e.g., time-domain SDNN, frequency-domain LF, HF and LF/HF ratio HRV, and non-linear metrics) will be tabulated consistent with best practices (Shaffer & Ginsberg, 2017).

### Mindfulness Intervention (MBSR)

We will adapt the MBSR intervention-authorized curriculum guide manual published at the Center for Mindfulness in Medicine, Health Care, and Society (CFM) at the University of Massachusetts Medical School (Santorelli et al., 2017). A systematic review (de Vibe et al., 2017) of the MBSR intervention examined over 100 RCTs, including over 8,000 participants, and concluded that MBSR has a moderately large effect on outcome measures of mental health, somatic health, and quality of life. The proposed adapted Mindfulness-Based Stress Reduction program will consist of 8 weekly 1 ½ hour sessions comprised of multiple components – guided meditations, breathe awareness, body scans (meditation to center awareness of bodily sensations and relieve tension), and guided mindful movements (adapted for the level of difficulty of participants). The sessions will be closed groups (because each session builds upon the previous session) with up to 10 participants per group and held on campus in a private meeting area. Evidence-based CM with up to approximately $120 in financial incentives for attending adapted MSBR sessions will occur to encourage retention. The final content and configuration of the intervention will be finalized based on feedback from our formative research.

### Assessment-only Condition

In the assessment-only condition, participants will undergo the same eligibility screening and assessments as those in the intervention group (see Figure 2). However, they will not participate in the 8-week adapted MBSR sessions. Instead, they will only complete the scheduled assessments at baseline, month 1, month 2, and month 3, and receive the same financial incentives for completing the assessments.

### Compensation Strategy

Participants will receive compensation for their involvement in the study to encourage retention and engagement. This includes payments for completing assessments (Baseline: $50, Post: $25, Follow-Up: $25), a qualitative post-intervention exit interview for Pilot trial participants ($25) and providing biomarker data ($10 for each timepoint).

### Language Considerations

Given that our previous work with Latine college students has shown a preference for English-language materials (Fernandez et al., 2019), the adapted MBSR program will be delivered in English. However, we will explore the possibility of offering Spanish-language cohorts in the future if needed.

### Staff Training and Quality Assurance

#### Training Procedures

Research staff and facilitators will undergo rigorous training to ensure the highest standards of research ethics, human subjects protection, and intervention delivery are met. Facilitators, selected for their experience with Latine populations and background in public health or social work, will receive specific training on the MBSR curriculum and its adaptations. Training will also cover all assessments and protocols.

#### Fidelity and Competence Assessment

All intervention sessions will be audiotaped, and facilitator performance will be evaluated against established competency criteria. Regular supervision meetings and feedback sessions will address any deviations from the protocol, ensuring consistent and high-quality intervention delivery.

#### Oversight and Monitoring

A Data Safety and Monitoring Plan (DSMP) structured in accordance with the National Institutes of Health (NIH) clinical trial guidelines will be implemented for the Cultural Adaptation Trial to ensure the safety of participants and the integrity of study procedures. The university’s Institutional Review Board (IRB) will provide additional oversight.

### Data Analysis Plan

#### Qualitative Analyses

All key informant interviews, focus groups, and pilot study exit interviews will be transcribed verbatim and coded using NVivo computer software. Transcriptions will be coded around primary content areas such as drinking and its consequences, mindfulness, program development, data collection, and recruitment. All transcripts will be independently analyzed by two coders whereby each coder will assign emergent themes through a descriptive, thematic analysis coding process. After each transcript is independently coded, coding teams will meet to review the reliability of code applications. Themes will then be created through a consensus process where coders reflect on emergent codes across all transcripts to develop overarching, broader themes. The objective of this process is to obtain relevant information to guide the adaptation, manual development, and finalization of the MBSR protocol intervention, data collection (self-report and biomarker) procedures, contingency management protocol, and dissemination strategies. This will be an interactive process, with data abstraction, coding, and categorization conducted simultaneously to identify salient themes and patterns.

#### Feasibility and Acceptability

Pilot and Cultural Adaptation Trial data will be analyzed for process indicators of feasibility such as % recruited who enroll; % who attend all sessions; % who complete post and follow up. The theoretical framework of acceptability (TFA) (Sekhon et al., 2017)will guide a multi-faceted assessment of acceptability, including ease of use (in daily life), relevance, and engagement, as well as satisfaction with intervention components and contingency management protocols. Proportions of individuals accepting, satisfied with, completing the intervention component, and retained in the study will be compared using chi-square tests. Additionally, we will use a priori minimum acceptability thresholds (e.g., > 80% retrospectively rating the intervention as acceptable at post-assessment).

#### Preliminary Assessment of Behavioral Outcomes

The main objective of the Cultural Adaptation Trial is to examine the acceptability and feasibility of the adapted MBSR program for Latine emerging adults engaging in problematic drinking, therefore, outcome analyses will be exploratory. Analyses will focus on within group changes over time and between group comparisons. Repeated measure analyses of covariance (ANCOVA) will be used to compare the study conditions on (a) retention, (b) problematic drinking and drinking-related consequences, and (c) putative mechanisms of change including reduced psychological stress, negative affect, greater self-regulation, and enhanced well-being. We will use effect sizes generated in this study along with participant feedback regarding usefulness to determine if further testing in a larger trial is indicated, although also acknowledge problems in the stability of effect size calculations with small samples. Additionally, we will explore more robust models of the associations of interest and possible mediator effects using structural equation modeling (SEM). Given the likelihood that drinking outcome variables will be non-normally distributed (e.g., zero-inflated, Poisson, negative-binomial), we will invoke various robust methods for addressing non-normality such as bootstrapping or applying a Hubert-White estimator as implemented in Mplus (Muthen & Muthen, 2007; Pek et al., 2018). To address any potential differences by sex, we will also test the heterogeneity of treatment effect (HTE) across biological sex by inserting an interaction term in the analytical model between group condition and sex. If significant, we will estimate differential effects of intervention within each sex subgroup. We expect the overall sex distribution to be 60%-40% with more females. As a result, we are powered to detect a medium to large intervention effect among females (d=0.67) and a large effect among males (d=0.83).

#### Sample Size and Statistical Power

Sample size justification, which is separate from a power calculation, appropriately considers the aims of the study and practical considerations of time and cost. The sample size planned here is consistent with the primary aims of assessing feasibility and use ecologically and will provide important initial data on main effects to inform a later clinical trial. However, to enhance our understanding of our ability to detect main effects with the Cultural Adaptation Trial (n=120), a priori power analysis was conducted using G*Power3 (Faul et al., 2007) for a repeated measures ANCOVA (between factors; 2 groups) using a two-tailed test, a medium effect size (f=.30), and an alpha level of .05. Results showed that a sample of 120 participants with two equal sized groups was sufficient to achieve a power of .80. Also, we plan to use SEM to explore mediator analyses, therefore sample size considerations are relevant for assessing the stability of the covariance matrices. Simulation studies (alpha level of 0.05 and a two-tailed test to achieve a minimum power of .80), suggest that a sample size of 120 is sufficient when using asymptotic theory for a wide range of latent variable models with well-defined factor structures (Jaccard & Wan, 1996; Jackson, 2003). The sample size in our Cultural Adaptation Trial meets this standard and will provide sufficient analytic power both within and across intervention conditions.

#### Data Management

During the start-up phase of the study, biomarker data collection procedures and quantitative and qualitative data programming, storage and management procedures will be established, including extensive data collection manuals and data logs. Custom surveys will be programmed and checked for accuracy using procedures developed in our previously funded studies. Database management, data visualizations and statistical analyses will be conducted throughout the project to ensure ongoing data monitoring and safety. Database, word processing, and graphics packages will be used, and statistical analyses will be performed using SPSS, SAS, R, Mplus, AMOS, and other relevant software.

#### Missing Data and Intent to Treat Analysis

In instances of missing data, values will be imputed using the Expectation-Maximization method with importance re-sampling as described in (King et al., 2001). If missing data are more substantial (e.g., > 12% of cases on a single variable or > 15% of cases have at least one missing value), multiple imputation will be used with five imputation data sets. We anticipate that the cultural and developmental adaptations and the CM component will address intrinsic and extrinsic participant motivation, engagement and retention. Still, some missing data due to attrition is likely. Thus, both intent-to-treat (priority) and dosage-response analyses will occur.

## Discussion

Emerging adults in the United States have the highest prevalence of alcohol use. While Latine emerging adults report lower rates of heavy alcohol use compared to their non-Latine White counterparts, those who drink face higher risks of transitioning to substance use disorders and experiencing more severe negative consequences. They are also less likely to seek treatment and have reduced access to innovative health-promoting resources, which exacerbates health inequities. Mindfulness-based interventions (MBIs) have shown promise in reducing problematic alcohol use and promoting healthier lifestyles, but existing research has limited minority representation and low adherence rates.

This study aims to decrease alcohol misuse among Latine emerging adults through a culturally adapted mindfulness-based intervention (MBSR). The overarching premise is that a culturally and developmentally tailored MBI can provide a respectful, non-judgmental, and discrimination-free environment that can appeal to Latine emerging adults who otherwise would not seek treatment for their alcohol misuse.

Using Community-Engaged Research (CEnR) methods, the study will adapt the intervention to be responsive to the target population’s needs. A pilot trial and a larger Cultural Adaptation Trial will evaluate the intervention’s acceptability, feasibility, and initial efficacy. To our knowledge, this will be the first study to adapt and evaluate the feasibility and acceptability of an empirically supported MBI on alcohol misuse among non-treatment-seeking Latine emerging adults. Further, the evaluation of embedding a contingency management (CM) approach to enhance MBI engagement and retention is novel. This study is an important step in the development of an effective intervention that can prevent escalation of alcohol misuse, and lead to improved health among this high-need population.

## Data Availability

not applicable. This manuscript only describes a planned protocol

## CRediT authorship contribution statement

Michelle M. Hospital: Conceptualization, Funding acquisition, Investigation, Methodology, Resources, Supervision, Validation, Writing – original draft, Writing – review & editing. María Eugenia Contreras-Pérez: Conceptualization, Methodology, Validation, Writing – original draft, Writing – review & editing. Sheila M. Alessi: Conceptualization, Funding acquisition, Investigation, Methodology, Resources, Supervision, Validation, Writing – review & editing. Staci Leon Morris: Funding acquisition, Writing – review & editing. Eric F. Wagner: Funding acquisition, Resources, Writing – review & editing. Robbert J. Langwerden, Augustin Castellanos, Katherine Perez & Jordan Quintana: Writing – review & editing.

